# Autoantibodies against the prion protein in individuals with *PRNP* mutations

**DOI:** 10.1101/19007773

**Authors:** Karl Frontzek, Manfredi Carta, Marco Losa, Mirka Epskamp, Georg Meisl, Alice Anane, Jean-Philippe Brandel, Ulrike Camenisch, Joaquín Castilla, Stéphane Haïk, Tuomas Knowles, Ewald Lindner, Andreas Lutterotti, Eric Vallabh Minikel, Ignazio Roiter, Jiri G. Safar, Raquel Sanchez-Valle, Dana Žáková, Simone Hornemann, Adriano Aguzzi

**Affiliations:** University of Zurich, Institute of Neuropathology, Zurich, Switzerland; University of Cambridge, Department of Chemistry, Cambridge, UK; CJD Foundation Israel, Pardes Hanna, Israel; ICM, Salpêtrière Hospital, Sorbonne University, Paris, France; University of Zurich, Institute of Surgical Pathology, Zurich, Switzerland; CIC bioGUNE and IKERBASQUE, Basque Foundation for Science, Bizkaia, Spain; Sorbonne University, ICM, Salpêtrière Hospital, Paris, France; University of Graz, Ophtalmology Division, Graz, Austria; University of Zurich, Department of Neurology, Neuroimmunology and MS Research (nims), Zürich, Switzerland; Broad Institute, Cambridge, USA; Treviso Hospital, Treviso, Italy; Case Western Reserve University, Department of Pathology, Neurology, and National Prion Disease Pathology Surveillance Center, Cleveland, USA; Alzheimer’s Disease and Other Cognitive Disorders Unit, Hospital Clinic, IDIBAPS, University of Barcelona, Barcelona, Spain; Slovak Medical University, Department of Prion Diseases, Bratislava, Slovakia

## Abstract

**Objective:** To determine whether naturally occurring autoantibodies against the prion protein are present in individuals with genetic prion disease mutations and controls, and if so, whether they are protective against prion disease.

**Methods:** In this case-control study, we collected 124 blood samples from individuals with a variety of pathogenic *PRNP* mutations and 78 control individuals with a positive family history of genetic prion disease but lacking disease-associated *PRNP* mutations. Antibody reactivity was measured using an indirect ELISA for the detection of human IgG_1-4_ antibodies against wild-type human prion protein. Multivariate linear regression models were constructed to analyze differences in autoantibody reactivity between a) *PRNP* mutation carriers versus controls and b) asymptomatic versus symptomatic *PRNP* mutation carriers. Robustness of results was examined in matched cohorts.

**Results:** We found that antibody reactivity was present in a subset of both *PRNP* mutation carriers and controls. Autoantibody levels were not influenced by *PRNP* mutation status nor clinical manifestation of prion disease. *Post hoc* analyses showed anti-PrP^C^ autoantibody titers to be independent of personal history of autoimmune disease and other immunological disorders, as well as *PRNP* codon 129 polymorphism.

**Conclusions:** Pathogenic *PRNP* variants do not notably stimulate antibody-mediated anti-PrP^C^ immunity. Anti-PrP^C^ IgG autoantibodies are not associated with the onset of prion disease. The presence of anti-PrP^C^ autoantibodies in the general population without any disease-specific association suggests that relatively high titers of naturally occurring antibodies are well tolerated. Clinicaltrials.gov identifier NCT02837705.

## Introduction

Prion diseases are diseases of the central nervous system which not only occur as sporadic and transmissible forms, but can also be transmitted through the germ line as autosomal dominant traits ^1^. Genetic prion diseases (gPrDs) account for ∼ 10-15 % of all prion diseases and are characterized by pathogenic, non-synonymous mutations of the human prion protein gene *PRNP* ^2^. The most prevalent human prion disease, sporadic Creutzfeldt-Jakob disease (sCJD), is characterized by a rapidly progressive dementia and a short survival time (usually < 1 year) from clinical onset ^3^. In contrast, *PRNP* mutation carriers often present with atypical phenotypes, e.g. long survival rates can be observed in Gerstmann-Sträussler-Scheinker disease (GSS) ^4^.

The cellular prion protein PrP^C^ consists of an unstructured, flexible tail (FT) on its N-terminal end and a C-terminal globular domain (GD) ^5^. We have shown in 2001 that humoral immunity against PrP^C^ can protect against prion neuroinvasion ^6^. Antibodies against the FT of PrP^C^, or removal of amino acid residues from the FT, abrogate the neurotoxic effects of anti-PrP^C^-GD antibodies and reduce the toxicity of *bona fide* prions ^7, 8^. Naturally occurring PrP antibodies may exist in the general population: for instance, reactivity against a 21-residue PrP peptide was observed in commercial pooled immunoglobulin ^9^, and a unique blood group has been observed in individuals homozygous for the E219K polymorphism ^10^.

Clinical trials have yet to deliver an effective anti-prion agent so far ^11-14^. An ongoing clinical study involves the administration of PRN100, a humanized antibody against PrP^C^-GD, to individuals suffering from CJD ^15^. While there is much hope that this trial will be successful, the murine counter-part of PRN100, ICSM18, exhibits an on-target, dose-dependent toxicity, and whether a therapeutic window exists has not yet been established ^16-18^.

The frequency of *PRNP* missense variants exceeds the reported genetic prion disease prevalence, suggesting a spectrum of disease penetrance in gPrDs rather than complete penetrance of non-synonymous *PRNP* mutations ^19^. The mechanisms by which these mutations induce disease are still largely unclear. The majority of structural studies on human PrP^C^ variants linked to genetic prion disease failed to identify consistent effects on global protein stability ^20^. Age of onset in gPrD is highly variable, and typically middle age or older, which might suggest that a protective mechanism guards some individuals against the prion protein-induced toxicity ^2^. We hypothesized that subtle conformational alterations of pathogenic PrP^C^ variants could stochastically generate immunogenic *neo*-epitopes, which in turn might elicit a protective humoral anti-PrP^C^ immune response. We therefore conducted an extensive search for such autoantibodies in individuals carrying pathogenic *PRNP* mutations, and in unaffected relatives as controls.

## Material and Methods

### Ethics statement

The Cantonal Ethics Committee of the Canton of Zurich approved this study (permit no. “KEK-ZH Nr.2015-0514”). This trial was registered at clinicaltrials.gov (no. NCT02837705). The protocol for this study was approved by the institutional review board at each participating institution with the University of Zurich being the lead regulatory site. Written informed patient consent was received by all individuals participating in this study.

### Human subjects and study design

We defined *PRNP* mutation carriers as individuals with a non-synonymous mutation in the open reading frame of the *PRNP* gene that was previously reported to be pathogenic ^2^. Between September 2015 and October 2018, we contacted both international patient organizations as well as national prion disease reference centers for further re-use of existing blood samples. Individuals at any age with a confirmed *PRNP* mutation were considered eligible for this study. Individuals with a confirmed *PRNP* mutation in a blood relative who did not undergo *PRNP* sequencing prior to enrollment in this study were also considered eligible if they gave consent for *PRNP* sequencing. Blood samples without information on age or gender were excluded from further analysis. *PRNP* wild-type individuals with neurological or psychiatric symptoms indicative of genetic prion disease were excluded from the study ^21^. Clinical manifestation of gPrD was defined as presence of both a pathogenic *PRNP* mutation and PrD-typical symptoms ^21^. The latter were assessed by clinical exam and neuropsychological assessment, in some cases complemented by ancillary tests such as presence of 14-3-3 proteins in cerebrospinal fluid, real-time quaking-induced conversion (RT-QuIC) assays, electroencephalography and magnetic resonance imaging ^22^. Personal history of autoimmune disease and other immunological disorders could be obtained in n = 141 of participants. A detailed description of the patient cohort is given in *table* 1. For sensitivity analysis, cases and controls were matched on age (± 5 years), gender and blood sample type (i.e. serum or plasma).

**table 1.** Baseline characteristics of the unmatched cohort.

|  | ***PRNP***<br>**mutation**<br>**carriers** | ***PRNP* wild-**<br>**type** | **Missing**<br>**Data**<br>**n (%)** | ***p* - Value** |
| --- | --- | --- | --- | --- |
| **Individuals enrolled,**<br>**n** | 124 | 78 |  |  |
| **Age** |  |  |  |  |
| **Mean (years)** | 49.3 | 42.8 |  | 0.004 |
| **SD (years)** | 16.5 | 13.9 |  |  |
| **Autoimmune**<br>**disease <sup>1</sup>** | 8 / 141 (5.7%) |  | 61 (30.2 %) |  |
| **Female gender,**<br>**n (%)** | 80 (64.5 %) | 37 (47.4 %) |  | 0.02 |
| **14-3-3 protein in**<br>**CSF** |  |  | 80 (39.6 %) <sup>2</sup> | n/a |
| **Test performed** | 17 / 63<br>(27.0 %) | 0 / 59<br>(0.0 %) |  |  |
| **Positive 14-3-3** | 8 / 17<br>(47.0 %) | n/a |  |  |
| **Codon 129**<br>**polymorphism, n (%)** |  |  | 20 (9.9 %) <sup>3</sup> | < 0.0001 |
| **Met / Met** | 69 / 121<br>(57.0 %) | 15 / 61<br>(24.6 %) |  |  |
| **Met / Val** | 50 / 121<br>(41.3 %) | 37 / 61<br>(60.7 %) |  |  |
| **Val / Val** | 2 / 121<br>(1.7 %) | 9 / 61<br>(14.8 %) |  |  |
| **Pathogenic *PRNP***<br>**mutation, n (%)** |  |  |  |  |
| **P102L** | 3 (2.4 %) | n/a |  |  |
| **D178N** | 37 (29.8 %) | n/a |  |  |
| **E200K** | 77 (62.1 %) | n/a |  |  |
| **V210I** | 2 (1.6 %) | n/a |  |  |
| **Unique <sup>4</sup>** | 5 (4.0 %) | n/a |  |  |

| Blood storage |  |  |
| --- | --- | --- |
| Plasma | 98 (79.0 %) | 70 (89.7 %) |
| Serum | 26 (21.0 %) | 8 (10.3 %) |

<sup>1</sup> due to few events of autoimmune disease, we pooled genotypes to eliminate possible identification. P-value compares numbers of individuals with or without autoimmune disease in *PRNP* mutation carriers and wild-type participants. <sup>2</sup> missing values: n = 61 *PRNP* mutation carriers, n = 19 *PRNP* wild-type. <sup>3</sup> missing values: n = 3 *PRNP* mutation carriers, n = 17 *PRNP* wild-type. <sup>4</sup> unique (n = 1) mutations: D178N/N171S, V180I, T183A, F198S, E200G.

### PRNP genotyping

*PRNP* genotyping was performed using a modified version of the DNeasy Blood & Tissue Kit (Qiagen). 20 μL of PK (600 mAU/ml) and 200 μL of 5 M guanidine hydrochloride (GdnHCl) with 1% Triton-X100 at pH = 5.0 were added to 200 μL of anticoagulated blood, vortexed thoroughly and incubated for 24 h at room temperature. 200 μL EtOH (96-100%) were added to the reaction and the rest of the DNA purification was performed according to the manufacturer’s guidelines. The primer pair PRNP_up and PRNP_low (*table e-1* available from Dryad https://doi.org/10.5061/dryad.08kprr4xk) was used in combination with Q5 high-fidelity DNA polymerase (New England Biolabs) to amplify the open reading frame from exon 2 of *PRNP*. Sanger sequencing was performed at the Department of Molecular Pathology (Institute of Surgical Pathology, University Hospital Zurich) using four different sequencing primers (PRNP_up, PRNP_up2, PRNP_low, PRNP_low2, *table e-1* available from Dryad https://doi.org/10.5061/dryad.08kprr4xk). Sequencing traces were aligned to reference DNA from the Reference Sequence (RefSeq) Database (O’Leary et al. 2016) using CLC Main Workbench (Qiagen) and packages sangerseqr^23^ and DECIPHER^24^ for Bioconductor^25^ in R.

### Statistical analyses

We performed *a priori* testing of anti-PrP^C^ autoantibody reactivity for the following hypotheses: a) differences in anti-PrP^C^ autoantibody reactivity between *PRNP* mutation carriers and *PRNP* wild-type individuals and b) differences in anti-PrP^C^ autoantibody reactivity between *PRNP* mutation carriers showing clinical signs of prion diseases and those without. All other analyses were performed *post hoc*. We used already established predictors of autoimmune disease such as age ^26^ and gender ^27^ as well as storage conditions known to affect antibody responses such as presence of coagulation factors ^28^ as covariates in our multivariate regression model. Using the *purposeful selection of covariates* method as described previously ^29^, effects of covariates on autoantibody titers were tested by bivariate linear regression analyses using the Wald test and included for multivariate testing at a p-value cut-off point of 0.25. In the multivariate model covariates were removed if they were non-significant at the 0.1 alpha level or not a confounder, as determined by a change in the remaining parameter estimate greater than 20 % as compared to the full model. *PRNP* mutation status, clinical signs of prion disease, *PRNP* codon 129 polymorphism were added after establishment of significant confounders. In matched cohorts, multivariate models were adjusted for matching factors.

All values are given as average ± standard deviation unless mentioned otherwise. For analysis, autoantibody titers were log_10_-transformed, and reported β coefficients and confidence intervals represent back-transformed values. Normality was tested using the D’Agostino-Pearson normality test. For values following a Gaussian distribution, differences between two groups were compared using two-tailed student’s T test. For not normally distributed values, Mann Whitney U Test was used for comparison of two groups. For comparison of categorical variables, Fisher’s exact test and chi-squared test were used for comparison of two and more than two groups, respectively. Pearson correlation coefficient was computed for data sampled from Gaussian distributions and Spearman’s rho for those sampled from non-Gaussian distributions. Matching of cases and controls was done using the *find*.*matches* function from the *Hmisc* package in R. We used lm for R for linear regression analysis. Python and R were used for statistical analysis, data visualization was performed using Prism 7 (GraphPad).

### Data availability statement

The study participants, if they have not undergone predictive testing themselves, participated under the condition of not knowing their *PRNP* genotype. Due to the relatively small sample size and risk of de-identification, all raw study data involving human participants was made available to the editors and reviewers but will not be made available publicly. Supplementary data, as well as DNA sequences of gene blocks used for construction of humanized antibodies and human PrP^C^-AviTag^™^ are available at Dryad https://doi.org/10.5061/dryad.08kprr4xk.

## Results

### Description of the cohort

We received blood samples and clinical information from a total of n = 241 individuals and selected n = 202 unmatched cases and controls for this analysis (*figure* 1). To test the robustness of our results, we matched n = 64 cases on n = 64 controls based on age (± 5 years), gender and blood storage conditions (i.e. serum / plasma, *table* e-2 available from Dryad https://doi.org/10.5061/dryad.08kprr4xk). Anti-PrP^C^ autoantibody reactivity was measured by a sandwich enzyme-linked immunosorbent assay (ELISA), a description of the assay is provided in *extended text* and *figures* e-1 and e-2 available from Dryad https://doi.org/10.5061/dryad.08kprr4xk. Briefly, blood samples were diluted over a range of > 2 logs and bound autoantibodies were detected with anti-human IgG antibodies. Antibody titers are expressed as negative common logarithm of the half-maximal effective concentration (*figure* e-1E available from Dryad https://doi.org/10.5061/dryad.08kprr4xk). Anti-PrP^C^ antibody reactivity was independent of serum IgG levels (Spearman’s ρ = 0.07, *p* = 0.69, *figure* 2A). The age of probands did not influence the IgG levels (Pearson r = 0.33, *p* = 0.16). To confirm our ability to detect human antibodies against specific targets, we tested a subset of individuals for the presence of IgG against the Epstein-Barr nuclear antigen (EBNA). 4/5 *PRNP*^WT^ and 16/16 *PRNP*^Mut^ individuals tested positive (corresponding to 95 % positive individuals), in line with anti-EBNA IgG seroprevalence in the general population (*figure* 2B) ^30^.

**Figure 1.**
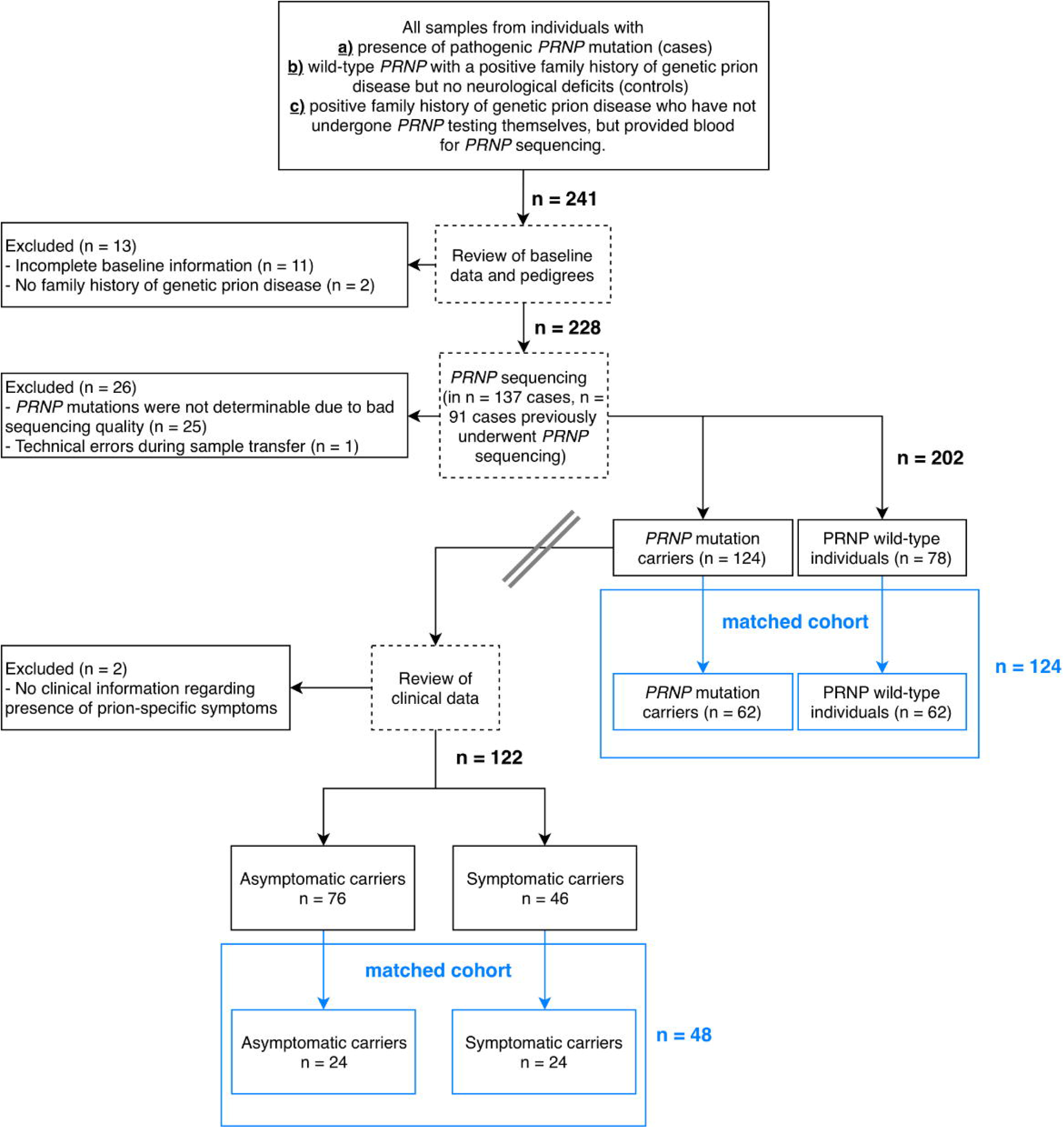
Flowchart of patient selection. Double line indicates cohorts selected for comparison of anti-PrP^C^ autoantibody titers from individuals carrying wild-type or mutated *PRNP* alleles (right of double line) and cohort selected for comparing anti-PrP^C^ autoantibody titers of symptomatic versus asymptomatic mutation carriers (left of double line). Blue boxes indicate matched cohorts.

**Figure 2.**
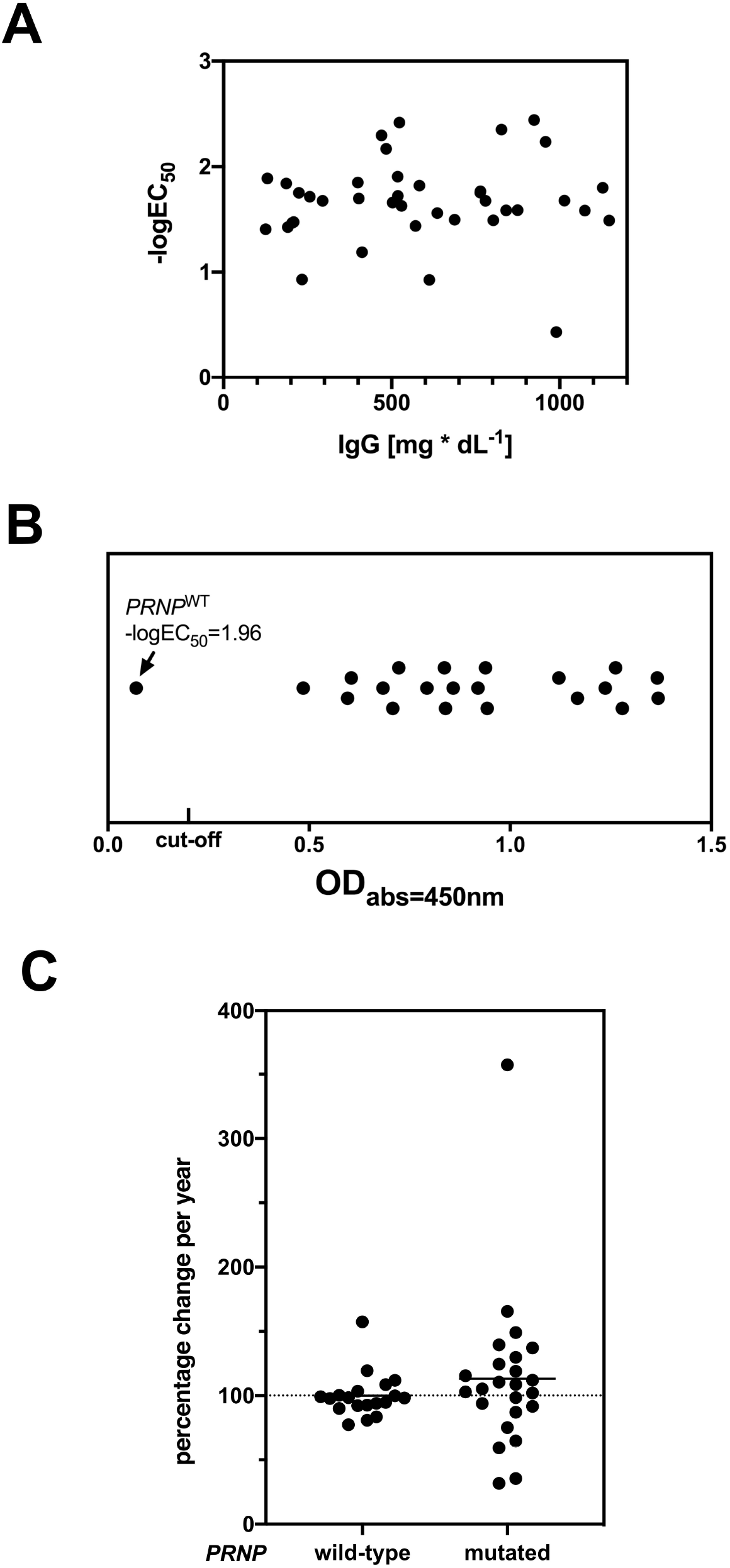
Correlation of anti-PrP^C^ autoantibody reactivity with total IgG levels, IgG anti-EBV autoantibodies and change of autoantibody titers over time. **(A)** Correlation of total IgG with anti-PrP^C^ autoantibody titers. **(B)** Qualitative assessment of anti-EBNA IgG antibodies in blood shows one *PRNP*^WT^ individual without detectable anti-EBNA IgG antibodies. *Cut-off*: OD_abs=450nm_ (optical density at absorbance α=450 nm) = 0.2 according to the manufacturer’s guidelines. **(C)** In two subsequent blood drawings, mean change of antibody titers per year is stable and similar between *PRNP* mutation and wild-type carriers, but variance is larger in *PRNP* mutation carriers.

### Prevalence of anti-PrP^C^ autoantibodies in PRNP mutation carriers

The presence of coagulation factors (e.g. plasma instead of serum), and possibly age, but not female gender were associated with anti-PrP^C^ autoantibody reactivity in bivariate and multivariate analyses (*table* 2) ^29^. We henceforth adjusted all analyses for age and presence of coagulation factors. Presence or absence of a pathogenic *PRNP* mutation was not associated with significant changes in anti-PrP^C^ autoantibody reactivity (*table* 3). Additionally, we matched both n = 62 cases and controls on age (± 5 years), gender and blood sample type ^26-28^ (*table* e-2 available from Dryad https://doi.org/10.5061/dryad.08kprr4xk). As with the unmatched cohort, *PRNP* mutation did not significantly influence anti-PrP^C^ autoantibody titers in multivariate linear regression adjusted for matching factors (*table* e-2).

**table 2.** Age and lack of coagulation factors in blood (e.g. serum probes), but not gender, are significantly associated with anti-PrP^C^ autoantibody reactivity. Due to lack of confounding effects of gender in multivariate model B, all further analyses were adjusted for blood sample type (serum/plasma) and age.

| **Bivariate analyses** |  |  |  |
| --- | --- | --- | --- |
| **Risk factor** | **β coefficient** | **95% confidence interval** | **p-Value** |
| **Age** | 0.990 | 0.982 - 0.998 | *0.05 \** |
| **Female gender** | 1.14 | 0.86 - 1.48 | 0.42 |
| **Plasma instead of serum** | 1.84 | 1.31 - 2.58 | *0.004 \** |
| **Multivariate Analysis - model A** |  |  |  |
| **Age** | 0.990 | 0.982 - 0.997 | *0.032 \** |
| **Plasma instead of serum** | 1.88 | 1.34 - 2.63 | *0.002 \** |
| **Multivariate Analysis - model B** |  |  |  |
| **Age** | 0.989 | 0.981 - 0.997 | *0.026 \** |
| **Female gender** | 1.16 | 0.90 - 1.50 | 0.35 |
| **Plasma instead of serum** | 1.86 | 1.33 - 2.61 | *0.003 \** |

**table 3.** Impact of *PRNP* mutation status on anti-PrP^C^ autoantibody reactivity.

| Risk factor | $\beta$ coefficient | 95% confidence interval | *p*-Value |
| --- | --- | --- | --- |
| ***PRNP* mutation (all)** |  |  |  |
| crude | 0.81 | 0.62 - 1.05 | 0.19 |
| adjusted | 0.92 | 0.70 - 1.20 | 0.61 |
| ***PRNP* D178N mutation** |  |  |  |
| crude | 0.61 | 0.44 - 0.86 | 0.02 * |
| adjusted | 0.75 | 0.53 - 1.06 | 0.17 |
| ***PRNP* E200K mutation** |  |  |  |
| crude | 1.16 | 0.89 - 1.16 | 0.36 |
| adjusted | 1.18 | 0.91 - 1.54 | 0.30 |
| **Clinical signs of prion disease** |  |  |  |
| crude | 0.90 | 0.59 - 1.38 | 0.64 |
| adjusted | 0.94 | 0.61 - 1.46 | 0.79 |

We then tested whether anti-PrP^C^ autoantibody response was associated with symptoms of prion disease. Presence or absence of clinical signs was reported by n = 122 *PRNP* mutation carriers (out of a total of n = 124 enrolled): n = 76 (62.3 %) were asymptomatic carriers whereas n = 46 (37.7%) presented with clinically apparent disease. Detailed clinical data was available in n = 14 cases, the most common clinical presentations entailed cerebellar signs (n = 12, 85.7%) and dementia (n = 11, 78.6%). Status of 14-3-3 protein in cerebrospinal fluid, albeit a poor predictor of genetic prion disease ^31^, was provided by n = 121 of study participants. N = 17 individuals (all *PRNP* mutation carriers with clinically apparent disease) were tested with n = 8 (47.1%) being tested positive, in line with previous findings ^31^. Presence of prion-specific symptoms was not associated with alterations in anti-PrP^C^ autoantibodies in an unmatched cohort (*table* 3). This was confirmed in an analysis of a cohort consisting of n = 24 symptomatic *PRNP* mutation carriers and n = 24 asymptomatic *PRNP* mutation carriers matched on *PRNP* mutation, age and sample type (*table* e-2).

### Post hoc subgroup analyses on the association of anti-PrP^C^ autoantibodies with specific PrP^C^ mutations, PrP^C^ p.129 polymorphism and autoimmune disease and other immunological disorders

We analyzed the effects of *PRNP* mutations that were present at least five times in the study population, namely D178N and E200K, on anti-PrP^C^ autoantibody titers: individuals with D178N mutations showed a significant trend towards lower autoantibody titers in bivariate analysis (*table* 3). This finding, however, was not significant after adjusting for age and sample type (*table* 3). E200K mutation carriers did not show significant changes in autoantibody reactivity (*table* 3). The (M)ethionine/(V)aline polymorphism at codon 129 of the human *PRNP* gene was reported to affect the susceptibility to prion diseases ^32^. Information on p.129 polymorphism was available in n = 182 of study participants: n = 84 (46.2%) were homozygous for methionine (p.129MM), n = 87 (47.8%) p.129MV, n = 11 (6.0%) p.129VV. None of the polymorphisms significantly altered autoantibody response to PrP^C^ in a *post hoc* analysis (*table* 4). In D178N carriers, the clinical phenotype was suggested to be dependent on the *PRNP cis* c.129 polymorphism: methionine was associated with FFI and valine with fCJD ^2^, although this association may not be universal ^33, 34^. In our cohort, n = 28 patients could be unambiguously identified as D178N_*cis*129M and n = 5 patients as D178N_*cis*129V. No differences in mean antibody reactivity were seen between those two groups (*table* 4).

Co-occurrence of multiple autoimmune diseases is a commonly observed phenomenon ^35^. In order to test the influence of pre-existing autoimmune diseases on anti-PrP^C^ autoantibody titers we searched clinical reports of study participants for presence of autoimmune disease and other immunological disorders. We were able to retrieve this information in n = 141 (69.8 %) of cases: n = 8 individuals were diagnosed with autoimmune disease, namely Hashimoto’s thyroiditis (n = 3), Graves’ disease (n = 1), monoclonal gammopathy of unknown significance (n = 1), multiple sclerosis (n = 1), psoriasis (n = 1) and rheumatoid arthritis (n = 1). Multivariate linear regression analysis adjusted for age, gender and type of blood sample did not show a significant association of autoimmune disease with anti-PrP^C^ autoantibody titers (*table 4*).

**table 4.** Impact of *PRNP* codon 129 polymorphism and history of autoimmune disease on anti-PrP^C^ autoantibody reactivity.

| Risk factor | $\beta$ coefficient | 95% confidence interval | *p*-Value |
| --- | --- | --- | --- |
| **p.129MM** |  |  |  |
| crude | 0.83 | 0.63 - 1.10 | 0.28 |
| adjusted | 0.88 | 0.67 - 1.15 | 0.43 |
| **p.129MV** |  |  |  |
| crude | 1.23 | 0.94 - 1.62 | 0.21 |
| adjusted | 1.13 | 0.86 - 1.48 | 0.46 |
| **p.129VV** |  |  |  |
| crude | 0.88 | 0.49 - 1.56 | 0.71 |
| adjusted | 1.04 | 0.59 - 1.83 | 0.90 |
| **D178N / cis-129M** |  |  |  |
| crude | 1.18 | 0.44 - 3.15 | 0.78 |
| adjusted | 1.09 | 0.37 - 3.22 | 0.90 |
| **History of autoimmune disease** |  |  |  |
| crude | 0.96 | 0.47 - 1.96 | 0.92 |
| adjusted | 1.12 | 0.55 - 2.26 | 0.76 |

### Temporal evolution of anti-PrP^C^ autoantibodies

44 individuals (21.8 %) donated blood multiple times, several months apart on which we performed a *post hoc*, time course analysis. *PRNP* wild-type individuals were observed over a longer time period compared to *PRNP* mutations carriers (17 ± 1.78 months vs. 10 ± 6.21 months, *p* = 1.42 × 10^−5^). *PRNP* mutation carriers showed larger variability in autoantibody titers, mean proportional change per year was, however, similar across groups (*p* = 0.23) and was overall negligible between two blood drawings (113.2 ± 61.44 % per year in *PRNP* mutation carriers versus 99.95 ± 17.22 % per year in *PRNP* wild-type individuals, *figure* 2C). None of the *PRNP* mutation carriers tested in this time course analysis exhibited clinical signs of prion disease.

## Discussion

The diagnosis of a disease-associated *PRNP* mutation is a fateful and often devastating event for individuals carrying such mutations. The clinical penetrance of *PRNP* mutations can be very high, and no disease-modifying therapy is available as of today ^2^. Clinical signs of familial prion disease typically erupt in late adulthood although carriers arguably produce the mutated protein from the first day of their life ^5^. There are at least two scenarios that may account for this phenomenon: (1) the pathogenic mutations may slightly destabilize PrP^C^, thereby infinitesimally increasing the probability of pathological aggregation, or (2) the pathogenic conformation of PrP^C^ is attained early on, but the body’s defenses stave off its consequences for many decades.

In the case of scenario #1, extensive structural studies on pathogenic PrP^C^ variants failed to reveal major structural alterations ^20^. We hypothesized that under scenario #2, the stochastic generation of PrP^Sc^ in mutation carriers might engender neoantigens, which in turn might lead to protective humoral responses. Remarkably, however, we found no evidence of induction of humoral antibody-mediated immunity against PrP^C^ by pathogenic *PRNP* variants. Instead, our study suggests the prevalence of naturally occurring anti-PrP^C^ antibodies in the general population independent of clinical signs of prion disease, *PRNP* variant or *PRNP* p.129 polymorphism. Although reactivity to wild-type PrP has been reported in the serum of E219K homozygotes ^10^, and reactivity to a non-naturally-occurring PrP peptide was reported in commercial IgG ^9^, the present report is to our knowledge the first observation of the *PRNP* genotype-independent presence of autoantibodies to full-length, wild-type PrP in humans. Without disease-specific antibodies, one might speculate that PRNP mutations accumulate subclinical levels of prions to a point when clinical symptoms become evident.

In a subset of individuals, anti-PrP^C^ autoantibody reactivity was tested in multiple blood drawings up to 1.5 years apart: the mean change of autoantibody titers was similar across *PRNP* genotypes in line with previous reports that showed stable autoantibody levels at least over several years ^36, 37^.

Matching in case-controls studies is a controversial topic ^38^. In our study, initial analyses were performed on unmatched cohorts adjusted for known confounders of blood autoantibody levels, this approach was described to increase statistical power ^39^. To strengthen our arguments, we compared anti-PrP^C^ autoantibody levels in cases and matched controls, these results are in line with findings from the unmatched cohorts.

An increasing number of autoantibodies against neurodegenerative targets are being explored as biomarkers and as potential therapeutics. Naturally occurring autoantibodies against hyperphosphorylated tau protein have been isolated from several asymptomatic blood donors ^40^. Researchers from Neurimmune (Switzerland) recently reported the development of a fully human antibody against amyotrophic lateral sclerosis targeting pathologically misfolded SOD1, α-miSOD1, from a memory B-cell library from healthy elderly individuals ^41^. Phase III trials involving aducanumab, a *bona fide* human antibody with potent β-amyloid clearing capabilities, were, however, stopped prematurely ^42^.

In previous works, we found that anti-PrP^C^ antibodies can efficaciously counteract prions ^6^ – a finding which was later confirmed by several other researchers ^43^. We speculated that anti-PrP^C^ autoantibodies from the general population could represent a reservoir of potential therapeutic agents against prion diseases. We find, however, that the distribution of titers appears similar between mutation carriers and controls, and between symptomatic and pre-symptomatic mutation carriers, arguing against the possibility that these autoantibodies are broadly beneficial. This is at variance with a previous, pre-clinical report claiming neuroprotective effects for naturally occurring antibodies to a PrP peptide ^9^. Similarly, naturally occurring anti-β-amyloid autoantibodies with neuroprotective effects were reported in mice, but did not meet primary cognitive endpoints when tested in a phase III clinical trial 44.

Nonetheless, our work does not rule out the possibility of protective anti-PrP autoantibodies in the general population or in *PRNP* mutation carriers specifically. Our study was restricted to the assessment of autoantibody levels against full-length, wild-type, recombinant human PrP^C^. We did not evaluate the presence of antibodies specific to pathogenic *PRNP* mutations or to neoepitopes created by those mutations. Moreover, it is possible that humans develop antibodies specific to PrP^Sc^, the aggregated form of the prion protein. In our experience such anti-PrP^Sc^ antibodies tend to cross-react, at least to some level, with PrP^C 45^. Another difficulty is that PrP^Sc^ structure is very heterogenous in genetic prion diseases: while brains from genetic CJD and sCJD patients show similar patterns of PrP^Sc^, PrP^Sc^ is fragmented and of low molecular weight in brains from GSS patients and can show marked variation in individuals with the D178N mutation ^2, 46^. Future studies will focus on the detection of rare, low-titer anti-PrP^Sc^ antibodies which may possess unique prion-clearing properties.

## Data Availability

The study participants, if they have not undergone predictive testing themselves, participated under the condition of not knowing their PRNP genotype. Due to the relatively small sample size and risk of de-identification, all raw study data involving human participants was made available to the editors and reviewers but will not be made available publicly.  DNA sequences of gene blocks used for construction of humanized antibodies and human PrPC-AviTagTM are available from the first author upon reasonable request.

## Acknowledgements

The authors wish to acknowledge their deepest gratitude to all individuals who participated in this study. The authors are impressed by the enthusiasm and generosity of the participating patients, which is a constant source of inspiration to perform biomedical research. The authors are grateful to the patients’ families, the CJD Foundation, referring clinicians, and all the members of the National Prion Disease Pathology Surveillance Center for invaluable technical help. The authors wish to thank Anne Kerschenmeyer, Tina Kottarathil and Rita Moos at the University Hospital of Zurich for excellent technical assistance.

The authors would like to thank the EPSRC, BBSRC, ERC and the Frances and Augustus Newman Foundation for financial support. This work was supported by the programs “Investissements d’avenir” ANR-10-IAIHU-06, “Santé Publique France” and supported by grants from NIH (R01NS103848), and CDC (UR8/CCU515004). Karl Frontzek received funding from the Theodor Ida Herzog-Egli Stifung and an unrestricted grant by Ono Pharmaceuticals. Georg Meisl is funded by a Ramon Jenkins Research Fellowship at Sidney Sussex College. Adriano Aguzzi is the recipient of an Advanced Grant of the European Research Council (ERC 250356) and is supported by grants from the Swiss National Foundation (SNF, including a Sinergia grant), the Swiss Initiative in Systems Biology, SystemsX.ch (PrionX, SynucleiX), the Klinische Forschungsschwerpunkte (KFSPs) “small RNAs” and “Human Hemato-Lymphatic Diseases”, and a Distinguished Investigator Award of the Nomis Foundation. Collection of samples at Massachusetts General Hospital was funded by Prion Alliance. The funders played no role in study design, data collection and analysis, decision to publish, or preparation of the manuscript.

**Appendix 1 – author’s contributions**

| Name | Location | Role | Contribution (see below) |
| --- | --- | --- | --- |
| Karl Frontzek,<br>MD PhD | University of Zurich,<br>Institute of Neuropathology,<br>Zurich, Switzerland | Author | Conceptualization, Data curation,<br>Formal analysis, Funding acquisition,<br>Investigation, Methodology, Project<br>administration, Software,<br>Supervision, Validation, Visualization,<br>Writing – original draft |
| Manfredi Carta,<br>MD | University of Zurich,<br>Institute of Neuropathology,<br>Zurich, Switzerland | Author | Investigation, Validation, Writing –<br>original draft |
| Marco Losa,<br>MD | University of Zurich,<br>Institute of Neuropathology,<br>Zurich, Switzerland | Author | Investigation, Validation, Writing –<br>original draft |
| Mirka<br>Epskamp, MSc | University of Zurich,<br>Institute of Neuropathology,<br>Zurich, Switzerland | Author | Investigation, Methodology,<br>Validation, Writing – original draft |
| Georg Meisl,<br>PhD | University of Cambridge,<br>Department of Chemistry,<br>Cambridge, UK | Author | Data curation, Formal analysis,<br>Software, Writing – original draft |
| Alice Anane,<br>ND | CJD Foundation Israel | Author | Data curation, Investigation,<br>Resources |
| Jean-Philippe<br>Brandel, MD | Sorbonne University, Paris,<br>France | Author | Data curation, Investigation,<br>Resources |
| Ulrike<br>Camenisch,<br>PhD | University of Zurich,<br>Institute of Surgical<br>Pathology,<br>Zurich, Switzerland | Author | Methodology, Resources, Writing –<br>review & editing |
| Joaquín<br>Castilla, PhD | CIC bioGUNE and<br>IKERBASQUE, Basque<br>Foundation for Science,<br>Bizkaia, Spain | Author | Data curation, Investigation,<br>Resources |
| Stéphane Haïk,<br>MD PhD | Sorbonne University, Paris,<br>France | Author | Data curation, Investigation,<br>Resources |
| Tuomas<br>Knowles, PhD | University of Cambridge,<br>Department of Chemistry,<br>Cambridge, UK | Author | Data curation, Formal analysis,<br>Software, Writing – review & editing |
| Ewald Lindner,<br>MD | University of Graz,<br>Ophtalmology Division,<br>Graz, Austria | Author | Data curation, Investigation,<br>Resources |
| Andreas<br>Lutterotti, MD | University of Zurich,<br>Department of Neurology,<br>Neuroimmunology and MS<br>Research (nims), Zürich,<br>Switzerland | Author | Methodology, Writing – review &<br>editing |
| Eric Vallabh<br>Minikel, PhD | Broad Institute, Cambridge,<br>USA | Author | Data curation, Investigation,<br>Resources, Writing – review & editing |
| Ignazio Roiter,<br>MD | Treviso Hospital, Treviso,<br>Italy | Author | Data curation, Investigation,<br>Resources |
| Jiri G. Safar,<br>MD | Case Western Reserve<br>University, Department of<br>Pathology, Neurology, and<br>National Prion Disease<br>Pathology Surveillance<br>Center, Cleveland, USA | Author | Data curation, Investigation,<br>Resources |
| Raquel<br>Sanchez-Valle,<br>MD PhD | Alzheimer's Disease and<br>Other Cognitive Disorders<br>Unit, Hospital Clinic,<br>IDIBAPS, University of<br>Barcelona, Barcelona, Spain | Author | Data curation, Investigation,<br>Resources |
| Dana Žáková,<br>PhD | Slovak Medical University,<br>Department of Prion<br>Diseases, Bratislava,<br>Slovakia | Author | Data curation, Investigation,<br>Resources |
| Simone<br>Hornemann,<br>PhD | University of Zurich,<br>Institute of Neuropathology,<br>Zurich, Switzerland | Author | Conceptualization, Supervision,<br>Writing – original draft |
| Adriano Aguzzi,<br>MD PhD | University of Zurich,<br>Institute of Neuropathology,<br>Zurich, Switzerland | Author | Conceptualization, Funding<br>acquisition, Methodology, Project<br>administration, Resources,<br>Supervision, Writing – original draft |

**Appendix 2 – Co-investigators**

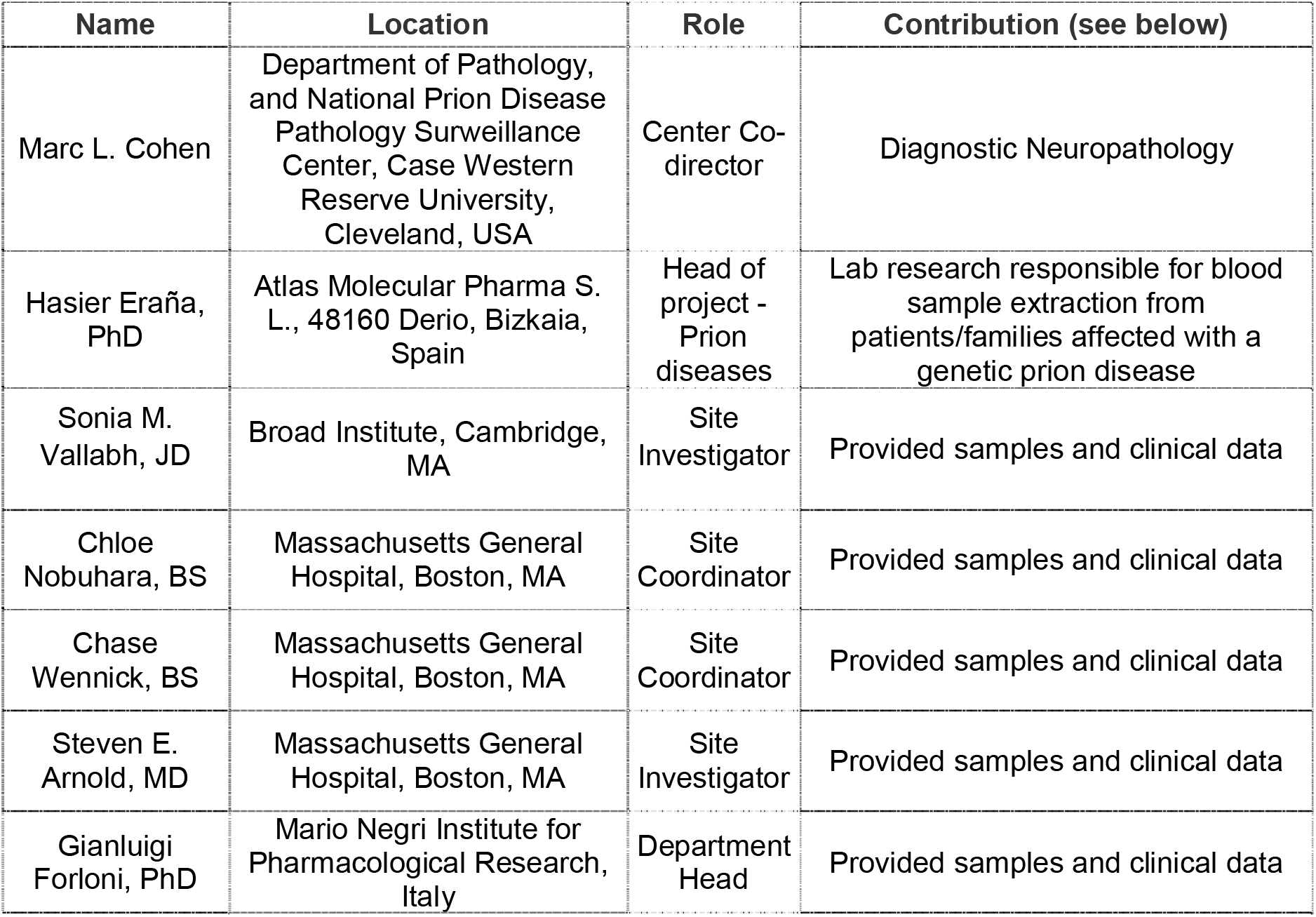

## Notes

**Author disclosures** Dr. Frontzek received an unrestricted grant by Ono Pharmaceuticals and was funded by the Theodor Ida Herzog-Egli Stiftung. Dr. Carta reports no disclosures. Dr. Losa reports no disclosures. Ms. Epskamp reports no disclosures. Dr. Meisl is funded by a Ramon Jenkins Research Fellowship at Sidney Sussex College. Ms. Anane reports no disclosures. Dr. Brandel reports no disclosures. Dr. Camenisch reports no disclosures. Dr. Castillas reports no disclosures. Dr. Roiter reports no disclosures. Dr. Haïk reports no disclosures. Dr. Minikel has received research support in the form of charitable contributions from Charles River Laboratories and Ionis Pharmaceuticals and has consulted for Deerfield Management. Dr. Knowles received financial support by the EPSRC, BBSRC, ERC and the Frances and Augustus Newman Foundation. Dr. Lindner was funded by the National Organization for Rare Diseases. Dr. Lutterotti reports no disclosures. Dr. Safar reports no disclosures. Dr. Sanchez-Valle reports no disclosures. Dr. Žáková reports no disclosures. Dr. Hornemann is the recipient of grants from SystemsX.ch (SynucleiX) and the innovations commission of the University Hospital of Zurich. Dr. Aguzzi is the recipient of an Advanced Grant of the European Research Council (ERC 250356) and is supported by grants from the Swiss National Foundation (SNF, including a Sinergia grant), the Swiss Initiative in Systems Biology, SystemsX.ch (PrionX, SynucleiX), the Klinische Forschungsschwerpunkte (KFSPs) “small RNAs” and “Human Hemato-Lymphatic Diseases”, and a Distinguished Investigator Award of the Nomis Foundation.

### Competing Interest Statement

Dr. Frontzek received an unrestricted grant by Ono Pharmaceuticals and was funded by the Theodor Ida Herzog-Egli Stiftung. 
Dr. Meisl is funded by a Ramon Jenkins Research Fellowship at Sidney Sussex College. 
Dr. Minikel has received research support in the form of charitable contributions from Charles River Laboratories and Ionis Pharmaceuticals and has consulted for Deerfield Management.
Dr. Knowles received financial support by the EPSRC, BBSRC, ERC and the Frances and Augustus Newman Foundation.
Dr. Lindner was funded by the National Organization for Rare Diseases.
Dr. Hornemann is the recipient of grants from SystemsX.ch (SynucleiX) and the innovations commission of the University Hospital of Zurich.
Dr. Aguzzi is the recipient of an Advanced Grant of the European Research Council (ERC 250356) and is supported by grants from the Swiss National Foundation (SNF, including a Sinergia grant), the Swiss Initiative in Systems Biology, SystemsX.ch (PrionX, SynucleiX), the Klinische Forschungsschwerpunkte (KFSPs) "small RNAs" and "Human Hemato-Lymphatic Diseases", and a Distinguished Investigator Award of the Nomis Foundation. 

### Clinical Trial

NCT02837705

### Author Declarations

All relevant ethical guidelines have been followed and any necessary IRB and/or ethics committee approvals have been obtained.

Any clinical trials involved have been registered with an ICMJE-approved registry such as ClinicalTrials.gov and the trial ID is included in the manuscript.

